# HMGB1 correlates with severity and death of COVID-19 patients

**DOI:** 10.1101/2022.05.26.22275611

**Authors:** Amanda Roberta Revoredo Vicentino, Vanderlei da Silva Fraga-Junior, Matheus Palazzo, Natalia Recardo Amorim Tasmo, Danielle A. S. Rodrigues, Shana Priscila Coutinho Barroso, Samila Natiane Ferreira, Anna Cristina Neves Borges, Diego Allonso, Marcelo Rosado Fantappié, Julio Scharfstein, Ana Carolina Oliveira, Rosane Vianna-Jorge, André Macedo Vale, Robson Coutinho-Silva, Luiz Eduardo Baggio Savio, Claudio Canetti, Claudia Farias Benjamim

## Abstract

SARS-CoV-2, the causative agent of the ongoing COVID-19, has spread worldwide since it was first identified in November 2019 in Wuhan. Since then, it was already demonstrated an exuberant inflammation, cytokine storm, endothelium dysfunction, platelets hyperactivation and aggregation, following T cell exhaustion leading to severe multi-organ damage and death of COVID-19 patients. Here, we sought to identify molecular biomarkers of disease severity in a Brazilian cohort of COVID-19 patients by measuring the serum levels of endogenous danger signals. Our data revealed that ICU patients that are critically ill, at the early hyperinflammatory phase of COVID-19 (around 12-25 days after hospital admission) display higher serum levels of the classical alarmin HMGB1. Serum levels of HMGB1 were positively correlated with cys-leukotrienes, D-dimer, AST, and ALT. Notably, we verified that HMGB1 levels above 125.4 ng/mL is the cut off that distinguishes the patients that are at higher risk of death. Serum levels of extracellular ATP, PGE_2_, LTB_4_, cys-LTs, and tissue factor were also elevated in the serum of ICU patients. In conclusion, we propose that serum levels of HMGB1 serve as prognostic biomarker of risk of death in patients suffering from severe COVID-19.

## INTRODUCTION

Since World Health Organization officially declared COVID-19 as a pandemic, unparallel efforts were dedicated to investigating the complex pathophysiology responses of the disease. Notwithstanding, the determinants of pathogenic outcome are still object of intense debate, and further studies are required to identify prognostic biomarkers and potential therapeutical targets. COVID-19 is caused by severe acute respiratory syndrome coronavirus 2 (SARS-CoV-2) and present a broad range of symptoms, ranging from asymptomatic and mild cases to severe and critical cases (10-20% of symptomatic patients; 15% progress to severe pneumonia, and about 5% are admitted to ICU due to acute respiratory distress syndrome [ARDS] and sepsis/multiple organ failure) (1). The world lethality rate is 2% accounting for more than 5 million deaths worldwide as of early January 2022. This new disease presents a very particular feature related to inflammatory mediators, cell migration and activation (1,2). In addition, long-term sequels have been described post-COVID-19 (3–7).

Patients with severe disease present high levels of IL-1β, IL-6, IP-10, TNF-α, reactive C protein (RCP), lactate dehydrogenase (LDH), and D-dimer, which are well-known markers of inflammation (8). An overwhelming immune response aggravates COVID-19 severity and worsens clinical outcome, eventually leading to death. Among the broad range of endogenous mediators of inflammation, high mobility group box 1 (HMGB1) is a classic example of an alarmin that is released to interstitial spaces following leukocyte activation or cell death-necrosis (9,10). HMGB1 has a major role in chronic inflammation, being implicated in inflammasome assembly (11,12), increased production of lipid mediators (13,14), and formation of neutrophil extracellular traps (NETS) following leukocyte infiltration in injured tissues (15).

It has been reported that HMGB1 levels in the blood of COVID-19 patients are increased, providing circumstantial evidence that this alarmin might contribute to disease outcome (16–18). Interestingly, HMGB1 increased angiotensin I-converting enzyme 2 (ACE2) expression on Calu-3, HepG2, Caco2, and RT4 cells via an end-product specific receptor (AGER). A different pathway,involving TLR4 signaling, is involved in TNF release by HMGB1 (17,19). Interestingly, elevated serum levels of S100A8/A9 and HMGB1 correlated with COVID-GRAM risk scores in critical illness in hospitalized patients (16). Extending the breadth of these studies, here measured the serum levels of extracellular ATP, PGE_2_, LTB_4_, cys-LTs, tissue factor (TF), and HMGB1 in COVID-19 severe patients, which were all elevated compared to health individuals. Based on our results, a cut-off HMGB1 serum level was defined above which it has been possible to predict a poor clinical prognosis and death for COVID-19 patients.

## SUBJECTS AND METHODS

### Study design and patient selection

This is a prospective study in collaboration with Biomedical Research Institute at Marcílio Dias Naval Hospital (Rio de Janeiro, Brazil). The sera from health donors and patients were collected in the period of March 2020 until December 2020, related to the first (SARS-CoV-2 Beta variant) and second waves (SARS-CoV-2 Gamma variant) of COVID-19 in Rio de Janeiro city (45) The patients were eligible if they tested positive for SARS-CoV-2 by reverse transcriptase-polymerase chain reaction (RT-PCR) for at least one respiratory sample. According to the sixth edition of Guidance for Corona Virus Disease 2019: Prevention, Control, Diagnosis and Management, issued by China’s National Health Commission, we only included in our cohort samples from severe COVID-19 patients admitted in the ICU with oxygen supplementation (93% intubated and 7% with face mask), pneumonia with ground-glass opacity, and high levels of D-dimer. We received samples from 73 patients randomly chosen from the HMMD repository of COVID-19 severe cases (322 total cases), with at least two samples collected in different hospitalization days. The period of hospitalization ranged between 6 and 186 days.. Thus, the data were analyzed considering the outcome and the respective stage of the disease on the date of blood collection. The forty-five healthy controls (HCs) consisted of volunteers’ participants recruited by public divulgation of the study. The HCs should not show any flu symptoms in the last 15 days before blood collection, and the participants who revealed detectable levels of anti-SARS-CoV-2 IgM or IgG were excluded from the cohort. Demographic data, comorbidities, clinical and biological parameters for COVID-19 patients were obtained from the medical record. For healthy controls these data were collected through anamnesis.

### Ethics statement

The present study was performed in accordance with regulations guidelines and approved by institutional ethical review boards from Clementino Fraga Filho University Hospital and Marcílio Dias Naval Hospital Ethics Committees (protocol numbers 4.551.702, protocol ID 361-20 and 32382820.3.0000.5256 respectively), with written informed consents obtained from all participants or their legal representatives.

### Nucleic acid extraction and cDNA synthesis

Total nucleic acid extractions from nasopharyngeal swab samples were performed using the automated Maxwell System platform (No AS4500, Promega, Madison, WI). Twenty-five microlitres of total RNA was submitted to cDNA synthesis with high-capacity cDNA reverse transcription kits (No 4368814, Thermo Fisher Scientific, Waltham, MA) and stored under - 20 °C. The extracted RNA was submitted to real-time RT-PCR for SARS-CoV-2 with of E and RP genes detection using the Fiocruz kit (Biomanguinhos, Rio de Janeiro, Brazil). Reverse transcription and amplification were performed in the QuantStudio 5 real-time PCR system (Thermo Fisher Scientific).

### Clinical samples and information collection

Blood samples were collected from hospitalized patients with severe COVID-19 (SARS-CoV-2-positive by RT–qPCR and serology) admitted to the intensive care unit (ICU) at Marcílio Dias Naval Hospital and HCs samples served as controls (SARS-CoV-2-negative by serology). Whole blood was collected in vacutainers and processed on the day of collection.

### Isolation of patient serum

Serum samples were collected after centrifugation of whole blood at 1200 g for 10 min at 4° C. The undiluted serum was then transferred to 0,5 mL polypropylene conical tubes, aliquoted, and stored at -80 °C for subsequent analysis. Repeated freeze-thaw cycles were avoided.

### ELISA for HMGB1

This assay was performed as described previously (46). Briefly, the wells of a 96-well microtiter plate (Greiner Bio-One, Austria) were coated overnight at 4 °C with anti-HMGB1 mouse monoclonal antibody (No H9537, Sigma-Aldrich, San Luis, MO) in PBS buffer (8.06 mM sodium phosphate, 1.94 mM potassium phosphate, 2.7 mM KCl, and 137 mM NaCl) at pH 7.4. The plates were blocked for 2 h at 37 °C with 1% BSA in PBS-T (0.05% Tween 20 in PBS) then washed five times with PBS-T buffer. Similar washing step was performed at the end of each incubation period. The wells were then incubated with serial dilutions of rHMGB1, ranging from 4,000 to 6.25 ng/mL, or with patients’ serum diluted 1:2 in PBS for 2 h at 37°C. Subsequently, the wells were incubated with rabbit-produced anti-rHMGB1 polyclonal antibody diluted in PBS buffer for 1 h at 37 °C, and then incubated for 1 h at the same temperature with anti-IgG rabbit antibody conjugated to horseradish peroxidase (No W4011, Promega, Madison, WI). The reactions were visualized with OPD (No P9187, Sigma-Aldrich) and H_2_O_2_ as the substrates and 12.5% H_2_SO_4_ as the quencher. Reactions were monitored by measuring the absorbance at 490 nm in a SpectraMax®M5/M5e Multimode Plate Reader (Molecular Devices, San José, CA). The standard curve calculation was performed using the mass value of the serial dilution of the rHMGB1 protein against its respective optical density (OD) measurement. The OD measurements were normalized using the mean value of the negative control replicates. Each patient sample was tested just once.

### Dosage of systemic levels of ATP

ATP circulating levels were measured in the serum of patients and healthy controls using a luciferase-based assay kit. The Molecular Probes® ATP Determination Kit (No A22066, Thermo Fisher Scientific) was used according to the manufacturer’s instructions. The luminescence of samples plated onto black 96-well plates was read in a SpectraMax®M5/M5e Multimode Plate Reader, and results were expressed as pmol of ATP.

### Measurement of lipid mediators

Lipid mediators present in the serum of healthy controls or SARS-CoV-2 infected patients were measured by commercial ELISA kits, according to the manufacturer’s instructions (PGE2 No. 514010, LTB_4_ No. 520111, and cys-LTs No.501070, Cayman Chemical, Ann Arbor, MI).

### Western blot analysis of tissue factor and transferrin

Serum from HCs and COVID-19 patients were reduced with β-mercaptoethanol in sample buffer. Samples were subjected to 10% polyacrylamide gel electrophoresis followed by transfer to nitrocellulose membrane. The immunoblot was performed using anti-tissue factor (1:1000, No ab104513, Abcam, Cambridge, United Kingdom) and anti-transferrin as load control (1:10000, No 82411, Abcam). The bands corresponding to both proteins were quantified using Image J software (NIH, Bethesda, MD) and the ratio between tissue factor and transferrin was calculated.

### Measurement of serum anti-SARS-CoV-2 antibodies

For quantitative analysis of anti-SARS-CoV-2 spike protein IgM and IgG antibodies, we performed the S-UFRJ test, as described previously (47). Briefly, high binding ELISA plates were coated with 50 μL of SARS-CoV-2 spike protein (4 μg/mL in PBS) and incubated overnight. The coating solution was removed and 100 μL of PBS 1% BSA (blocking solution) was added and the plate was incubated at room temperature for 1-2 h. The blocking solution was removed and 50 μL of diluted 1:40 (PBS 1% BSA) patient sera were added, subsequently, incubated at RT for 2 h. Then, the plate was washed with 150 μL of PBS (5x) and 50 μL of 1:10000 goat anti-human IgM and IgG (Fc)-horseradish peroxidase antibody (Sourthen Biotech, Birmingham, AL) were added, and the plate was incubated for 1.5 h at RT. The plate was washed again with 150 μL of PBS (5x) and then treated with TMB (3,3’, 5,5; -tetramethylbenzidine - Scienco, Brazil) until the reaction was stopped with 50 μL of HCl 1N. The optical density (OD) was read at 450 nm with 655 nm background compensation in a microplate reader (Bio-Rad Laboratories, Inc, CA).

### Statistical analysis

A descriptive study of the cohort was conducted, presenting measures of central tendency and dispersion for continuous variables, or relative frequencies for each categorical variable. The Kolmogorov-Smirnov test was employed to assess whether the variables were normally distributed. Continuous variables were presented as means ± standard deviations if normally distributed or as medians with 95% confidence interval (95% CI) interquartile ranges (IQRs) if not normally distributed. Categorical variables were defined according to better- or worse-expected prognostic values. Continuous variables were compared among multiple groups using ANOVA followed by the Student-Newman-Keuls test for normally distributed variables or the Kruskal–Wallis test if variables were not normally distributed. The non-parametric Mann–Whitney U test was used for univariate comparisons of selected variables according to primary outcomes (discharged *vs* diseased). The ability of HMGB to predict the final clinical outcome was analyzed using the receiving operating characteristic (ROC) curve and the area under the curve (AUC), accompanied by the 95% confidence interval. The optimal cut-off value was determined by the point of maximal sensitivity and specificity, and then used to calculate the associate relative risk (RR) and 95% CI of the worse outcome (death). All P-values reported are from two-sided tests. The threshold for significance was set at P = 0.05 and highlighted in bold in Tables and figures. Statistical analyses were performed in R version 3.1.1.

## RESULTS

### Cohort characteristics

COVID-19 subjects (n=61) and HC (n=45) were recruited between March 23^rd^ and December 03^rd^, 2020, and blood samples were collected and screened for several parameters. We received samples from 73 patients, nevertheless we excluded 12 participants because they acquired COVID-19 after hospitalization for other reasons such as cancer, risk pregnancy, car accident, knife or gun accident or absence of medical records. Sixty-one COVID-19 patients admitted to the intensive care unit (ICU) after 1 to 14 days of symptom onset participated in this study in which 57 needed invasive ventilation and 4 used oxygen supplementation. Seven patients denied comorbidities. COVID-19 patients were tested positive for SARS-CoV-2 by RT-PCR (Figure 1A). HC did not differ from patients related to age (COVID-19= 28-86 years old, mean= 62.8 years old; HC= 28-79 years old, mean= 65 years old), gender, and comorbidity frequencies (Figure 1B-C). As expected, our COVID-19 cohort was mainly composed of males (Figure 1B), corroborating a greater probability of men progressing to a more serious outcome (1,2). The cohort of healthy controls and COVID-19 patients presented the same comorbidities although in different frequencies. The most common underlying diseases in our cohort were hypertension (patient=72.1% vs HC=33.3%), obesity (patient=44.3% vs HC=17.8%), and diabetes (patient=29.5% vs HC=13.3%). It is worthy to state that a significant number of patients presented two or more comorbidities (Figure 1C). The comorbidity frequencies of hypertension, diabetes, chronic lung disease and vascular disease showed significant differences between HC and COVID-19 cohorts while comorbidity frequencies of obesity and chronic heart disease were not statistic different (Figure 1B-C). The COVID-19 subjects included in our cohort were categorized as severe/critical patients since all were in ICU, presented pneumonia and the need for mechanical ventilation (93% received invasive ventilation, intubation, and 7% received oxygen mask support), whereas 60.7% patients deceased. We also observed a mean of 6 days of symptoms onset until hospital admission and 38 days of hospitalization, similar data to those observed in the literature (20,21; Figure 1D). The symptoms reported by our COVID-19 patients agreed with other studies with a predominance of fever, cough, dyspnea, fatigue, and myalgia (Figure 1E).

**Figure 1.**
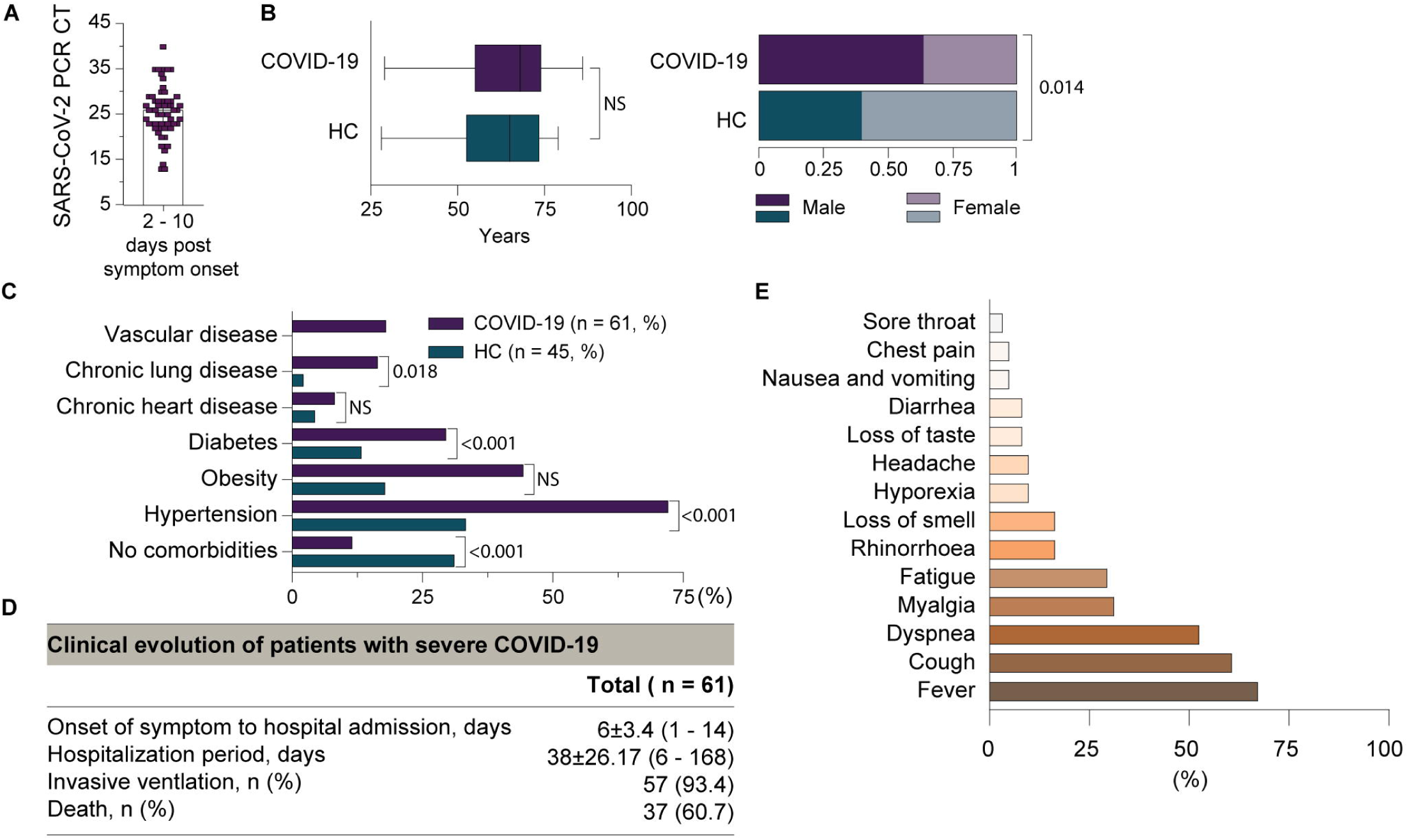
Demographic and clinical characteristics of patients during hospitalization and healthy controls. (A) SARS-CoV-2 PCR cycle threshold (CT) of nasopharyngeal swab after symptom onset of patients’ admission in Marcílio Dias Naval Hospital. (B) The age and gender are shown for the two different study groups. (C) Chronic comorbidities frequency within HC and COVID-19 patients. (D) Clinical characteristics as period of symptoms onset, hospitalization period, severity, and primary outcome of COVID-19 patients. Results are presented as mean ± standard deviation and range. (E) Main presenting symptoms of COVID-19 patients. Non-parametric t-test was used to analyze the age. CHI-square online was used to analyze the frequency of gender and comorbidities. P < 0.05 was considered statistically significant compared to HC; ns—not significant.

The humoral response of COVID-19 patients was evaluated, and as illustrated, SARS-CoV-2 infection caused an enhancement of anti-S protein IgM and IgG levels above the threshold limit (OD ratio above 2; Figure 2A-B), besides the positive PCR test (Figure 1A). The HC included in our work, in addition to the statement of not having contracted COVID-19, presented low levels (OD ratio under 2) of IgM and IgG anti-S protein. HC presenting OD ratio above 2 were excluded from the cohort. Interestingly, we observed some COVID-19 patients with low levels of anti-S antibodies. This finding was already observed in other studies and probably it is due to poor antibody producer individuals as elders which have low humoral response. The quantitative antibody response was also evaluated among discharged and deceased severe COVID-19 patients. The antibody levels were reduced, despite not being statistically different, in patients with fatal outcome compared to discharged patients, but it is still higher than non-COVID-19 subjects (Figure 2C-D - bottom graphs). Of note, the few COVID-19 patients with low levels of IgM and IgG (values under the OD ratio, panel C and D) deceased, suggesting that they were unable to present humoral response due to their critical ill condition.

**Figure 2.**
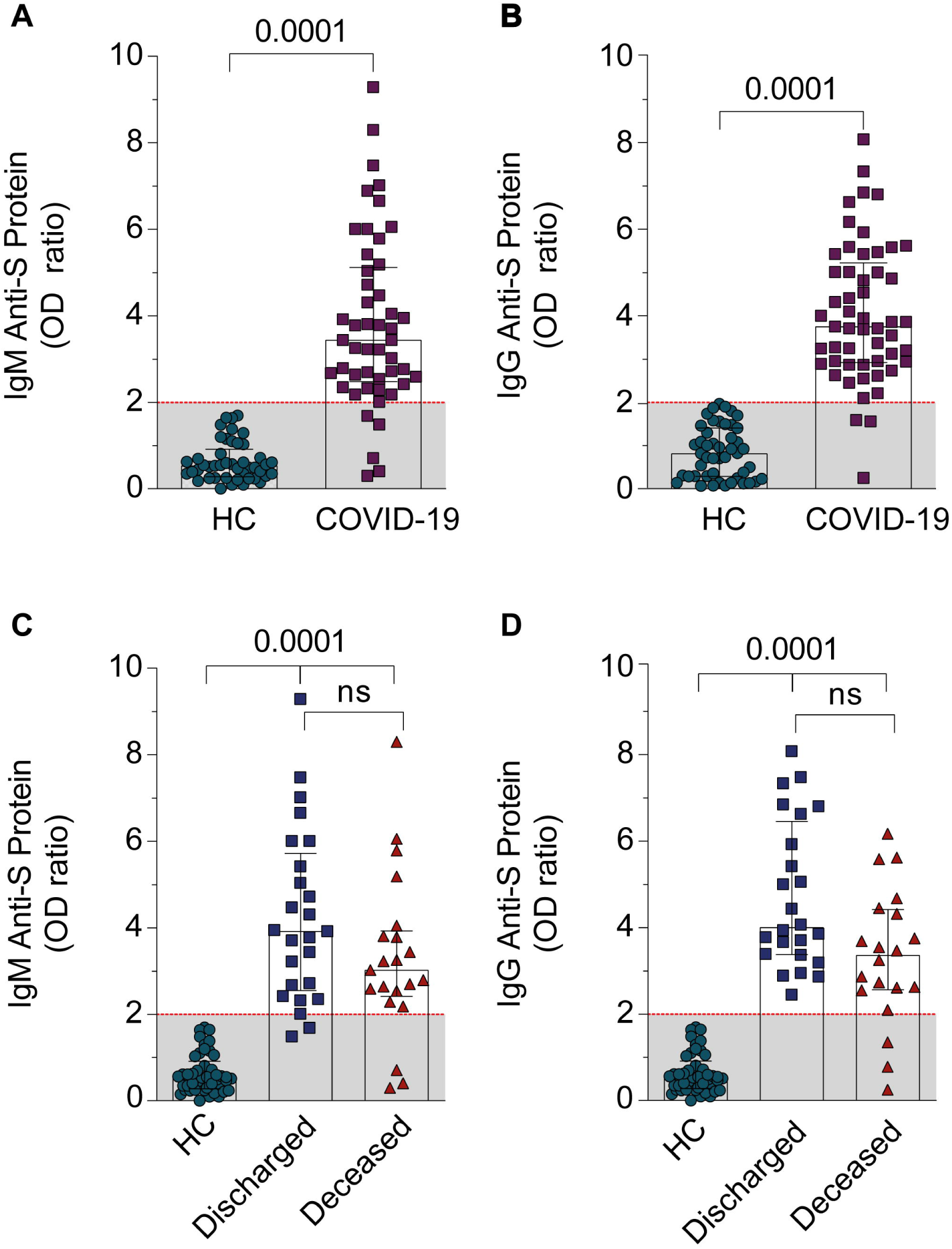
Anti-S protein immunoglobulin profile of COVID-19 patients and healthy controls. (A-B) Serum IgM and IgG levels of HC and COVID-19 groups. We plotted the highest value obtained after analysis of two or three samples received from each COVID-19 patient. (C-D) Serum IgM and IgG levels from discharged or deceased patients. The data were presented as the median with interquartile range and were analyzed by Mann-Whitney test (B, C) and Kruskal-Wallis test (D, E). P < 0.05 was considered statistically significant compared to HC; ns—not significant.

### COVID-19 patients present altered biochemical parameters

To further confirm the systemic commitment of COVID-19 patients and to evaluate correlations with the target inflammatory mediators, we analyzed the data of several organ dysfunction markers from medical records between discharged and deceased patients. The values of the COVID-19 subjects’ cohort were compared to the reference values detached as dashed red lines. One limitation was the fact that all COVID-19 subjects were critically ill, thus most biochemical markers of severity were similar between discharged and deceased patients (Figure 3). Nevertheless, deceased COVID-19 patients presented reduced number of platelets and PO_2_/FIO_2_ and increased lactate and CRP levels, compared with discharged patients (Figure 3B, I, J and L). About the lactate levels, we didn’t find levels above reference levels probably because both groups of patients were been finely controlling to avoid the lactic acidosis.

**Figure 3.**
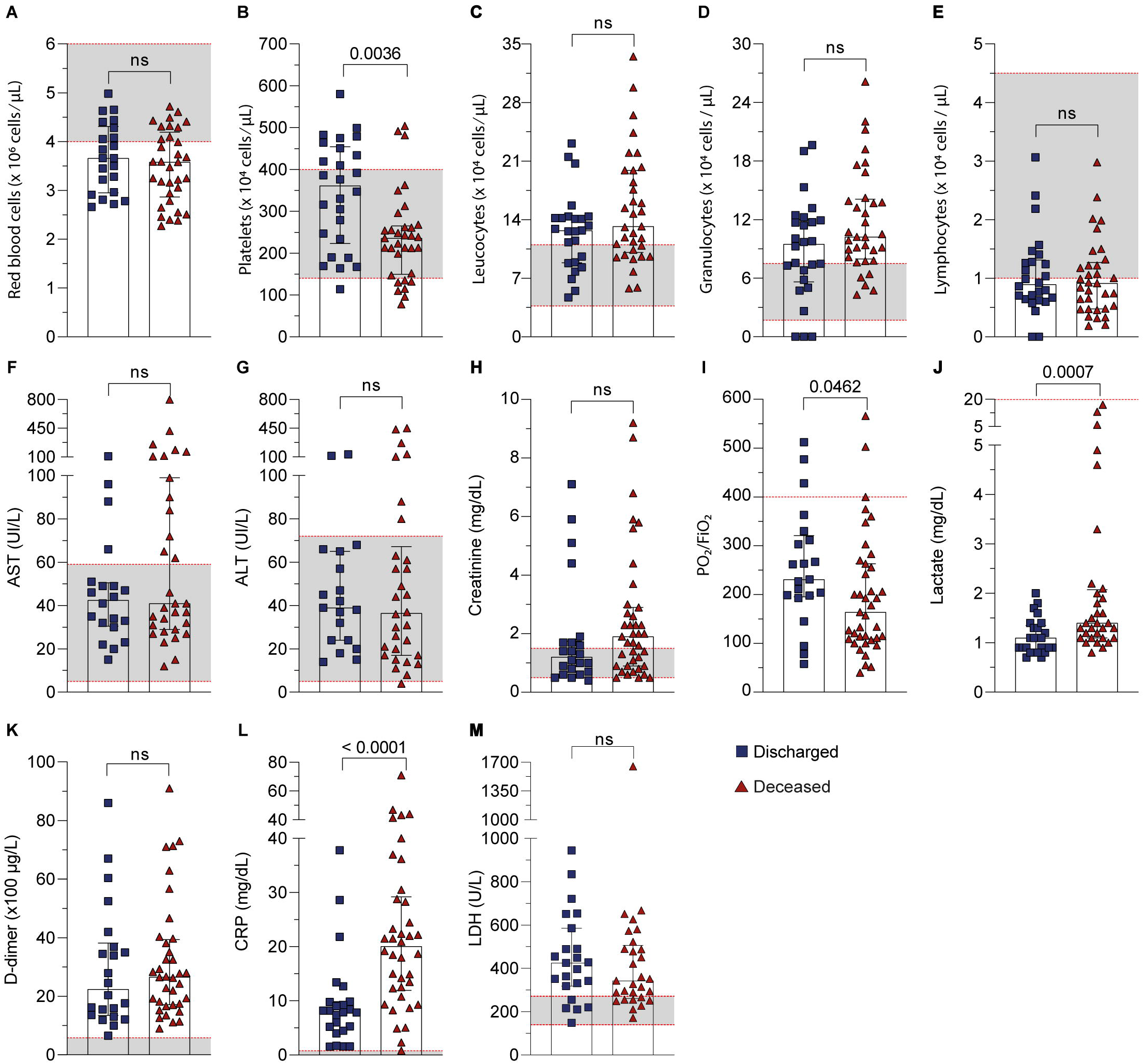
COVID-19 patients display alterations in biochemical parameters according to clinical outcome. Routine laboratory values in the circulation of discharged and deceased COVID-19 patients. (A-E) Number of red blood cells, platelets, and white blood cells. (F-I) Serum levels of hepatic (AST and ALT), kidney (creatinine) and lung (P/F) injury biomarkers. (J-M) Systemic inflammatory biomarkers (lactate, D-dimer, CRP, LDH). The range of references values for all parameters was marked as dashed red line. The data were presented as the median with interquartile range and were analyzed by Mann-Whitney test. P < 0.05 was considered statistically significant compared to discharged patients’ group. ns - not significant; AST - aspartate aminotransferase; ALT - alanine aminotransferase; CRP – C reactive protein; LDH – lactate dehydrogenase.

Regarding other systemic parameters, although they were not significantly different between groups, the number of red blood cells and lymphocytes were below the reference values, while total leukocytes and granulocytes were above (Figure 3A, C, D, and E). AST, ALT, and creatinine levels were in the normal range in most patients, having some patients with higher levels (Figure 3F, G, and H). D-dimer and LDH levels of both discharged and deceased patients were way over the reference levels (Figure 3K-M).

### Increase inflammatory mediators in COVID-19 patients

Inflammatory storm is well described and is one of the main causes of tissue damage and worst prognosis of COVID-19 (8). Among several mediators involved, we demonstrated that alarmins as HMGB1 and ATP were significantly increased in the serum of COVID-19 patients, compared to HC (Figure 4A-B). In order to characterize vascular activation, we quantified TF in the serum of HC and COVID-19 patients. We also detected higher levels of TF in COVID-19 patients compared to HC (Figure 4C, uncut gels). Lipid mediators play important roles in acute and late immune response during sepsis (22). Thus, we also demonstrated increased levels of PGE_2_, LTB_4_, and cys-LTs in the serum of severe COVID-19 patients (Figure 4D-F).

**Figure 4.**
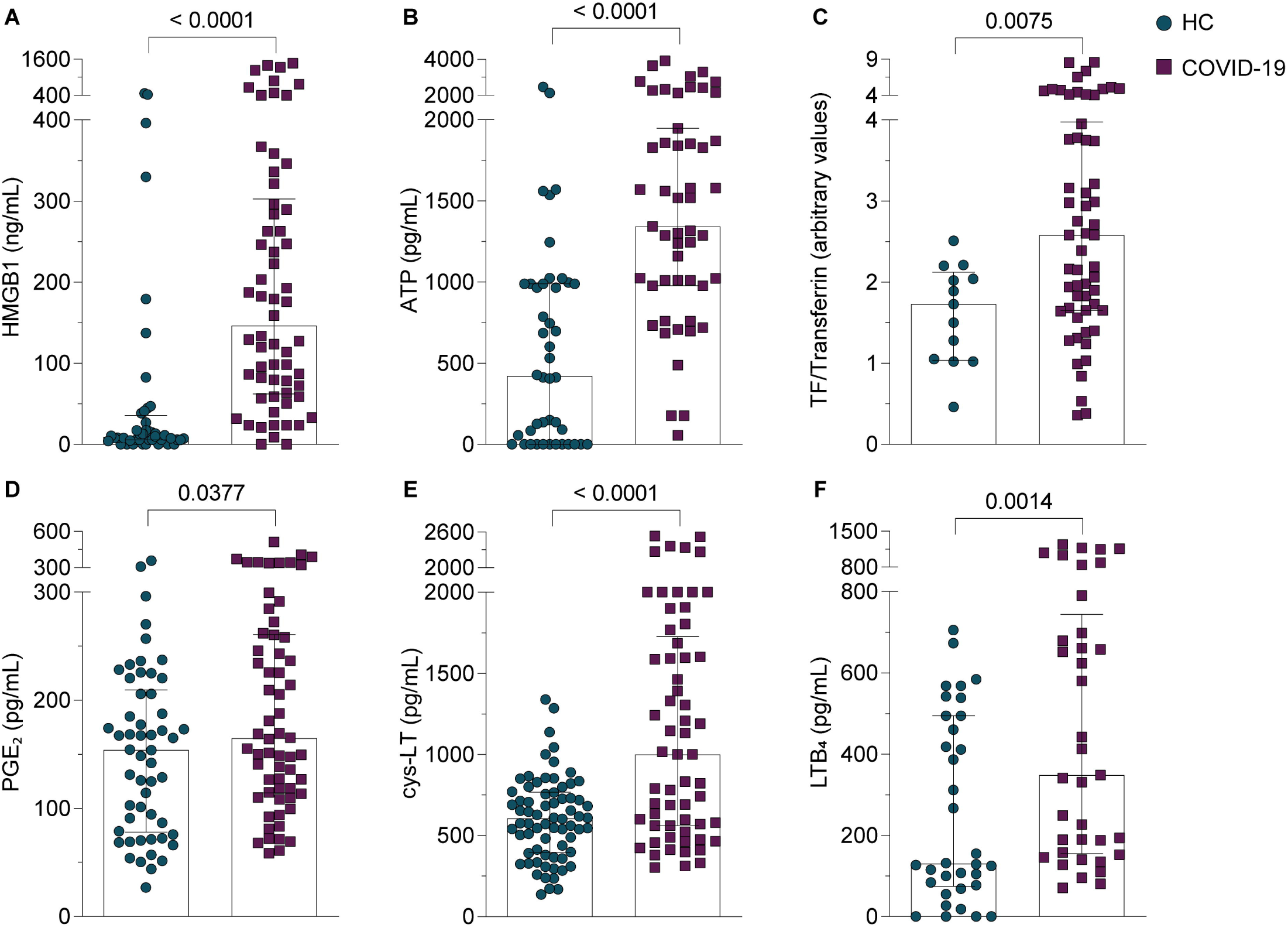
COVID-19 patients present high serum levels of HMGB1, ATP, TF, PGE_2_, cys-LT, and LTB_4_ compared to healthy controls. (A-F) We plotted the highest serum value obtained after analysis of two or three samples received from each COVID-19 patient. Mediators were evaluated as described in Methods section. Data were presented as the median with interquartile range and were analyzed by Mann-Whitney test. P < 0.05 was considered statistically significant compared to HC.

Further, we correlated those mediators with the primary outcome. HMGB1 was the only mediator showing higher levels in the deceased group compared to discharged group (Figure 5A). Even though we could not find statistical differences of cys-LTs levels between discharged and deceased patients; apparently, patients with fatal outcomes presented higher levels of this lipid mediator; however, the number of patients was not big enough to demonstrate statistical differences (Figure 5E). No differences were noted between discharged and deceased patients regarding to serum ATP, TF, PGE_2_, and LTB_4_ levels (Figure 5B-D, and F).

**Figure 5.**
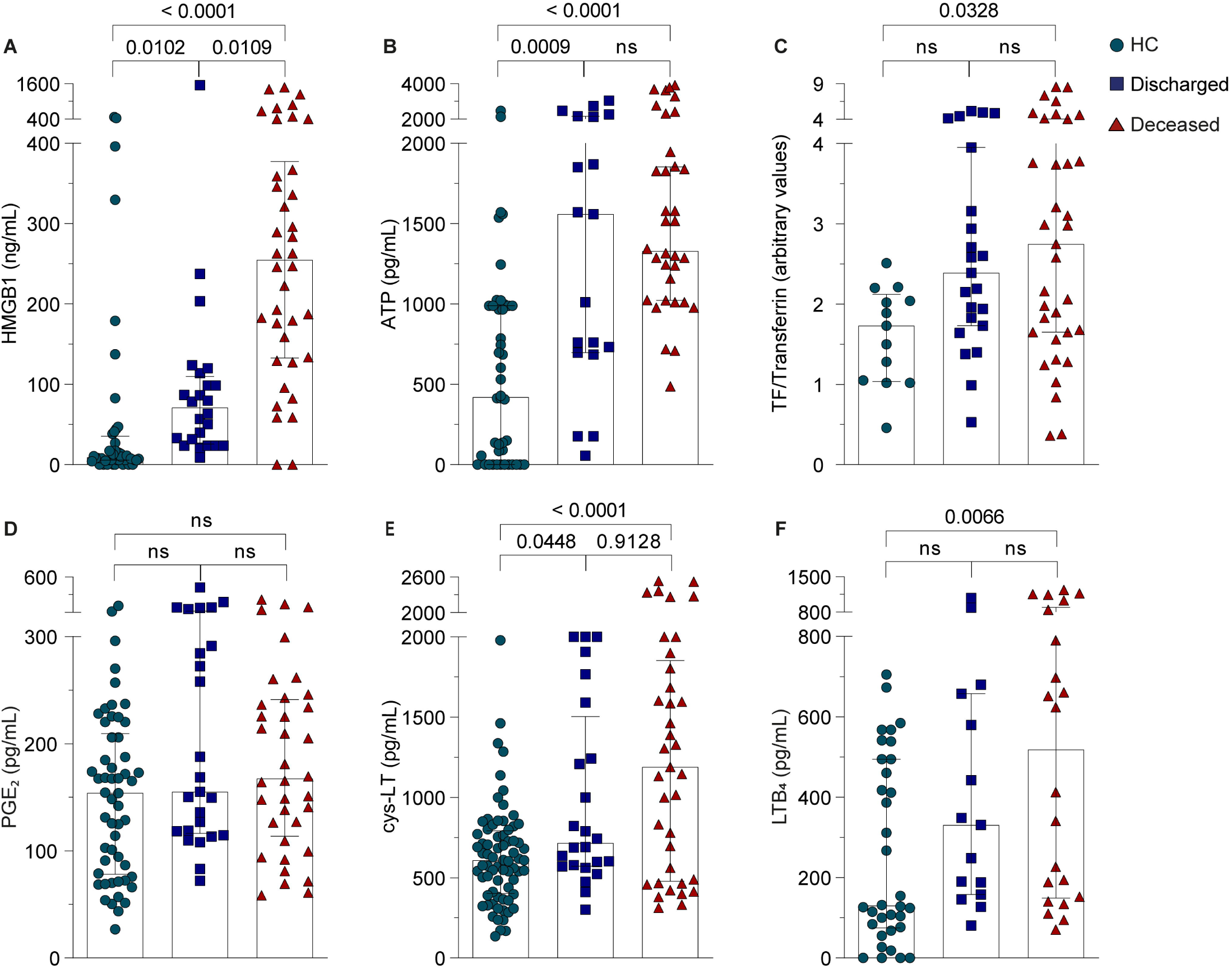
HMGB1 serum levels discriminate deceased from discharged COVID-19 patients. (A-F) HMGB1, ATP, TF, PGE_2_, cys-LT, and LTB_4_ levels were stratified between discharged and deceased in COVID-19 patients and compared to healthy controls. We plotted the highest serum value obtained after analysis of two or three samples received from each COVID-19 patient. The data were presented as the median with interquartile range and were analyzed by Kruskal-Wallis test. ns – not significant. P < 0.05 was considered statistically significant compared to HC or discharged groups.

### HMBG1 is higher in deceased versus discharged COVID-19 patients in the pulmonary phase with hypoxia and in the hyperinflammatory early phase

Looking for a specific marker that could predict the worst prognosis, we stratified the COVID-19 patients by different time phases of disease evolution: 1. pulmonary phase with hypoxia - from onset of the disease until day 11; 2. Hyper inflammatory early phase - period of 12-25 days; and 3. Hyperinflammatory late phase - period of 26-108 days. These phases were already described in the literature (20,21). HMGB1 was the sole mediator tested with significantly higher levels in the serum of patients who died compared with discharged patients during the pulmonary phase with hypoxia and in the hyper inflammatory early phase (Figure 6A). None of the other mediators analyzed showed differences.

**Figure 6.**
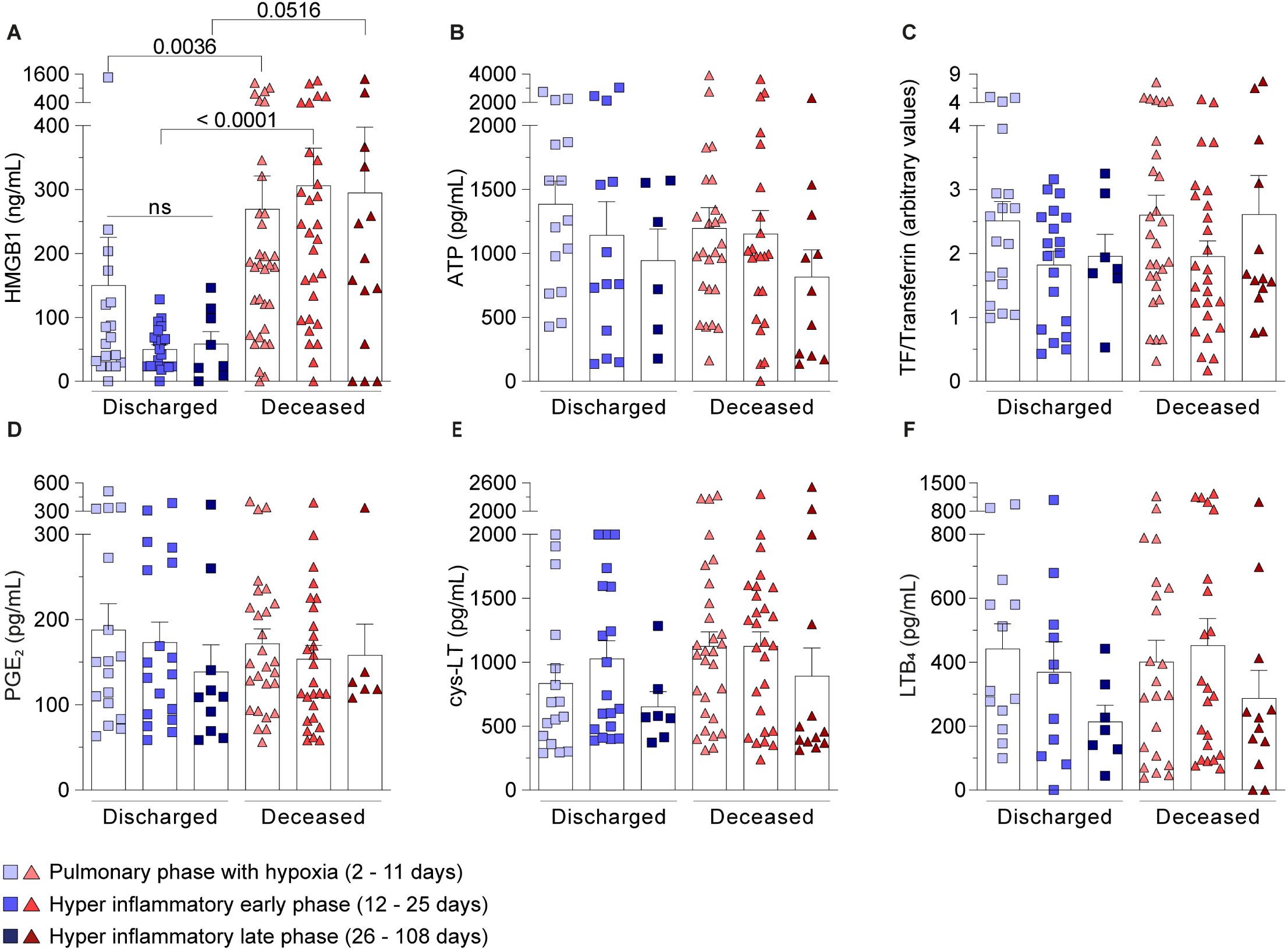
Higher HMGB1 serum levels were observed in the pulmonary phase with hypoxia and hyper inflammatory early phase according to clinical outcome. (A-F) The mediator values were stratified among three temporal phases after hospital admission: pulmonary phase with hypoxia (2-11 days), hyper inflammatory early phase (12-25 days), and hyper inflammatory late phase (26-108 days). Data were presented as mean ± SEM and were analyzed by Kruskal-Wallis test. ns – not significant. P < 0.05 was considered statistically significant compared to the indicate phases.

### HMGB1 predicts disease progression

We analyzed the correlation between serum levels of inflammatory mediators and the biochemical data obtained from patient records. Interestingly, we observed a significant correlation between HMGB1 and cys-LT (Figure 7A). Furthermore, HMGB1 also correlates with AST and ALT levels (Figure 7B-C), hepatic dysfunction markers. A positive correlation trend between HMGB1 and D-dimer was observed but did not reach significance (P=0.09; Figure 7D). We also analyzed correlation with several other markers in our study, but we could not find statistical differences (Supplementary Figure 1).

**Figure 7.**
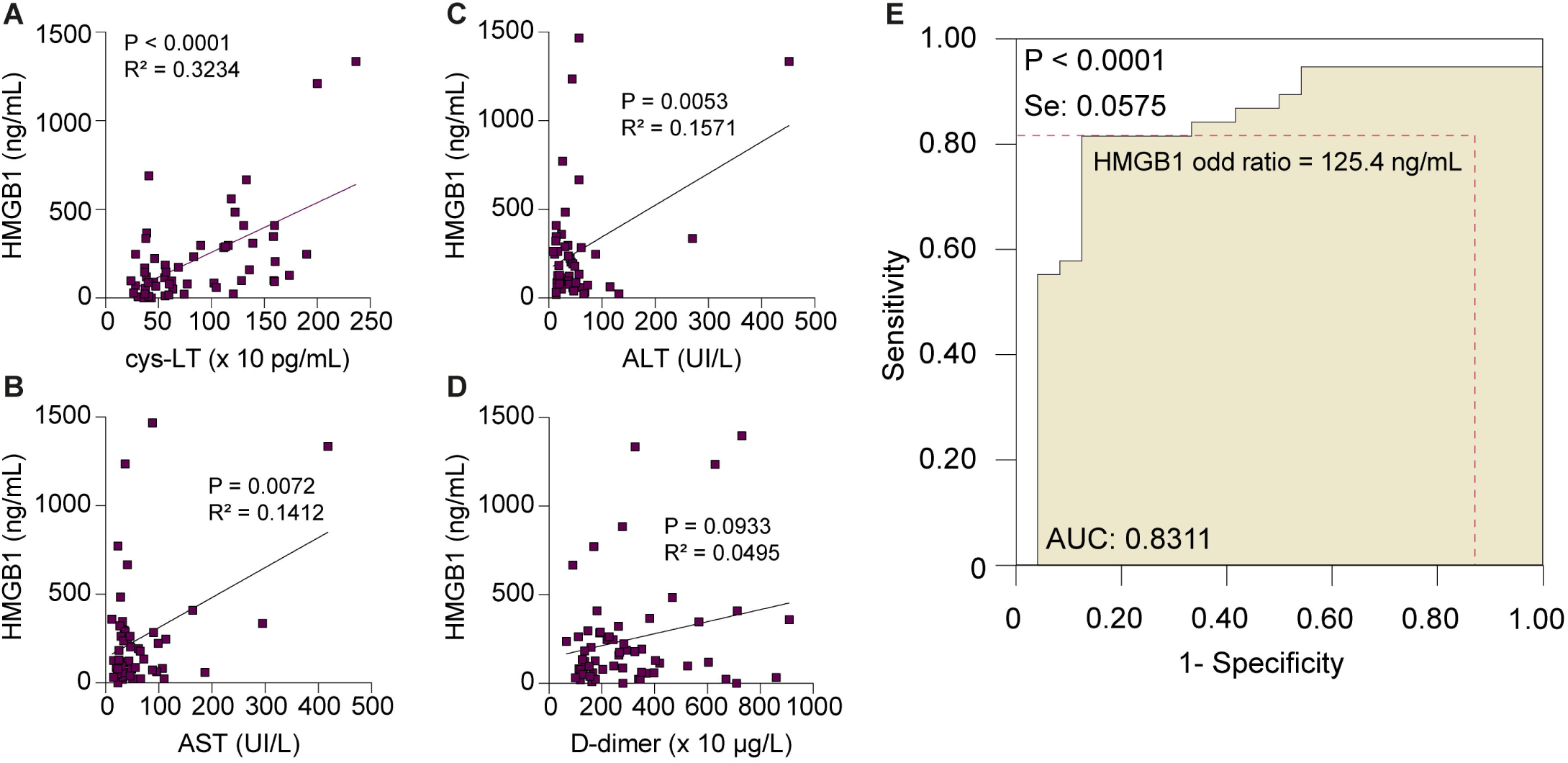
Positive correlations of HMGB1 with cys-LTs, AST, ALT, and D-dimer in COVID-19 patients. (A-D) Spearman’s correlation analyses between HMGB1 serum levels peak versus cys-LTs, AST, ALT, and D-dimer levels. We plotted the highest value obtained for HMGB1 after analysis of two or three samples received from each COVID-19 patient. The values for cys-LT, AST, ALT and D-dimer were paired exactly with the same sample chosen for analysis of HMGB1. (E) ROC curve analysis determines the HMGB1 cutoff value (125.4 ng/mL) that predicts the worst clinical outcome (death). The AUC is 0.8311 ± 0.06 and the 95% confidence interval (CI) is 0.7185 to 0.9437. The diagnostic sensitivity and specificity are 81.6 % and 87.5 %, respective

To explore the exact impact of HMGB1 levels on the disease progression, we examined the ROC curves with our data. The ROC cutoff value of serum HMGB1 levels that distinguishes discharged and deceased patients was 125.4 ng/mL. The diagnostic sensitivity and specificity for deceased patients was 81.6 % and 87.5 %, respectively (Figure 7E). These data demonstrated that values above 125.4 ng/mL predicted disease progression and worst outcome. This finding suggested that serum HMGB1 levels until the 12^nd^ day after hospital admission could be regarded as a biomarker for distinguishing COVID-19 patient outcomes.

## DISCUSSION

The acute phase of COVID-19 is characterized by a variety of immune alterations associated with endothelium dysfunction and uncontrolled inflammatory responses, including lymphopenia and cytokine storm mainly in the severe cases, affecting tissue integrity (1,2,8,23). Intriguingly, increasing evidence suggest that phenotypic and functional alterations of the immune system persist for a long period after recovery from COVID-19 (24,25).

A wide range of host immune responses is triggered by SARS-CoV-2, from appropriate and protective to uncontrolled and highly dysfunctional reactions. In this report we provided evidence of high levels of HMGB1, ATP, TF, cys-LTs, PGE_2_, and LTB_4_ in the circulation of severe COVID-19 patients. Still, patients who died from COVID-19 showed higher serum levels of HMGB1, compared to discharged patients, fact that leads us to consider HMGB1 a biomarker of fatality risk in a cohort of hospitalized patients.

The innate immune response, as the first line of defense against the infection, plays an outsized role in promoting inflammatory dysfunction, changing metabolic processes, and promoting malfunction of the adaptative immune response, such as T cells exhaustion and inadequate B cells/antibody response (1,26). HMGB1 and ATP stand out among the alarmins increased during COVID-19 infection (16–18,27). Although the pathogenic role of HMGB1 in COVID-19 remains to be explored in animal models, studies in culture systems demonstrated that HMGB1 up-regulates ACE2 expression, the main SARS-CoV-2 gateway on cellular membrane (17,19). Interestingly, ATP may favor increased levels of HMGB1 during COVID-19, since ATP triggers HMGB1 release by P2X7 receptor activation in a monosodium urate crystals-induced model of sterile inflammation (28). In the same line, eicosanoids such as cys-LTs, PGE_2_, and LTB_4_ are also released by ATP and HMGB1 stimuli (14,29–31). In agreement, some data in the literature demonstrated that lipid mediators were increased in severe COVID-19 patients and may be associated with poor outcomes (23,32). Meanwhile, the role of eicosanoids in COVID-19 remains to be characterized. Our data place the lipid mediators, mainly the cys-LTs, as an interesting target in COVID-19 in association with ATP and HMGB1. As well described, COVID-19 is a multi-mediated disease, and we attempt to characterize not just one mediator but the main scenario responsible for promoting disease.

Molecular analysis revealed that HMGB1 forms complexes with LPS or IL-1β to enhance immune responses (33–35) and ACE2 expression (17,19). While admitting that clinical studies cannot provide conclusive answers to this question, there is urgent need to identify markers of disease severity in COVID-19. Following the first report that HMGB1 levels were increased in severe COVID-19 patients, another study suggested that serum HMGB1 and IL-6 high levels may serve as prognostic biomarkers, distinguishing patients with unfavorable clinical outcome (18). Along similar lines, the serum levels of ATP and activation of CD39/CD73 axis may aggravate COVID-19 by harnessing cytokine production, inflammasome activation, cell death, and tissue damage (36).

The detection of high TF levels in the serum of severe COVID-19 patients raises the possibility that HMGB1 might be implicated in TF-driven hyperactivation of platelets, perhaps contributing to the thromboembolic complications observed in severe cases. Our cohort studies confirmed that cases of severe COVID-19 presented high levels of HMGB1, D-dimers, CRP, low ratio of PO2/FiO2 and lymphopenia. At first sight, these findings suggest that HMGB1, presumably acting as a pleiotropic driver of inflammation, may potentiate TF-dependent activation of platelets and circulating monocytes (37).

Interestingly, HMGB1 levels were able to discriminate between discharge and deceased patients. The cys-LTs levels also seemed to be higher in deceased patients, however with the stratification of patients for the primary outcome, the statistical analysis loses strength due to the small number of patients remained per group. Extending this analysis, we generated data during the evolution of patient’s hospitalization period. We divided this period into three phases of COVID development based on inflammatory parameters and clinical reports: 1) pulmonary phase with hypoxia (2-11 days after hospital admission); 2) hyper inflammatory early phase (12-25 days after hospital admission); and 3) hyper inflammatory late phase (26-108 days after hospital admission). Notably, HMGB1 was the sole cytokine with high serum levels during the pulmonary phase with hypoxia and during the hyper inflammatory early phase of patients who died. The implication of this finding must be emphasized because HMGB1 levels measured at the early inflammatory phase of the disease has predictive value for pathogenic outcome. This may be extremely valuable at the timepoint in which patients are hospitalized showing signs of cytokine storm. Our findings suggest that serum values of HMGB1 above 125.4 ng/mL in critically ill COVID-19 patients correlate with the worst outcome and that those patients may be at higher risk of death. Thus, measurements of serum HMGB1 levels may help to instruct pharmacological and medical interventions at early stage of disease. Importantly, as described in the analysis of this cohort, CRP levels were the unique clinical parameter that was found in higher levels in patients who deceased. So, HMGB1 as a severity biomarker of COVID-19 could open a new strategy for reverting ongoing processes skewing the innate and acquired immune response to an efficient and pro-resolution mode, instead of overwhelming inflammation and immune dysfunction.

In our work, we also evaluated the correlation between HMGB1 serum levels and several biochemical and inflammatory mediators and found a positive correlation with levels of cys-LTs. Reminiscent of sepsis-like inflammatory dysfunctions, it is possible that the HMGB1-cys-LT axis is involved in endothelial cell activation and long-term cytokine production by activated leukocytes, leading to systemic complications. A positive correlation between HMGB1 and ALT/AST was expected since our group already demonstrated that the liver is one of the main sources of systemic HMGB1 during infection (38). It could explain the hepatic dysfunction and alteration in immune metabolism. High levels of D-dimer have been reported in moderate and severe COVID-19 patients (39). All patients in our cohort presented high levels of D-dimer, regardless of whether they were discharged or died. The correlations with ATP, TF, PGE_2_, LTB_4_, creatinine, PO_2_/FiO_2_, lactate, CRP and LDH also did not reach statistical significance; further experiments are necessary to better understand the relationship of those mediators and systemic dysfunction markers.

Remarkably, our study demonstrates a relevant role of HMGB1 in the COVID-19 evolution. This cytokine stimulates the innate system either by itself or in association with endogenous and exogenous molecules (43). Another host molecule that acts as alarmin includes S100A8/A9, which is also released from dead cells as HMGB1 and ATP. Overproduced S100A8/A9 and HMGB1 in serum of COVID-19 patients were associated with distinct signatures for cytokine storm, and both are poor prognostic indicators (16). In parallel, we also believe in a partnership between HMGB1 and hypoxia-inducible factor-1α (HIF-1α) favoring the pathogenesis of COVID-19. As previously demonstrated (44), HIF-1α and HMGB1 are released by monocyte and endothelial cells, respectively, under hypoxic conditions observed during COVID-19. However, while HMGB1 promotes inflammatory cytokine production, elevated levels of HIF-1α repress IRF3 and IRF5 leading to low levels of type I IFN. Those mediators (S100A8/A9 and HIF-1α) may act in synergism with HMGB1, and these aspects are under investigation in our clinical trial and using animal model.

In conclusion, this scenario could explain the pathophysiology initiated by SARS-CoV-2 and place HMGB1 as one of the major mediators in triggering an overwhelming inflammatory cascade which impairs the host to wipe out the virus and exerts important deleterious effects in tissues and organs. Therefore, our work suggests that high levels of HMGB1 in the circulation of severe COVID-19 patients orchestrates the acute and persistent mediators’ storm, in association with several other mediators, which certainly prompts to the long-term consequences already well described.

The association of HMGB1 and COVID-19 has already been documented before by two independent group. Despite that, our work adds and ratifies the importance of this mediator as a promising biomarker in Brazilian cohort. Thus, our study is relevant to validate HMGB1 as a severity biomarker not only for COVID-19 but also for other diseases that present hyperinflammatory response. In addition, our study also analyzed ATP, cys-LTs, PGE_2_, LTB_4_ and TF which appear to act collaboratively to form an inflammatory platform, process that are under investigation to better understand the molecular mechanism behind COVID-19 pathophysiology.

## STUDY HIGHLIGHTS

### What is the current knowledge on the topic?

Pneumonia and acute respiratory distress syndrome are the major complications of COVID-19. SARS-CoV-2 infection can activate innate and adaptive immune responses and result in massive inflammatory responses later in the disease. These uncontrolled inflammatory responses may lead to local and systemic tissue damage and co-infections because of IUC environment. This scenario difficult the direct treatment of the disease. It is necessary the investigation of biomarkers that precociously distinguish the worst prognosis.

### What question did this study address?

In this study we present the levels of endogenous danger signals as HMGB1, ATP, TF, PGE2 in a Brazilian cohort of severe COVID-19 patients.

### What does this study add to our knowledge?

In this work we found that the levels of endogenous danger signals were altered as well as biochemical indicators, but only HMGB1 had distinguished values of serum levels between discharged and deceased patients in pulmonary and early hyperinflammatory phase of severe COVID-19.

### How might this change clinical pharmacology or translational science?

The results suggest that the alarmin HMGB1 could be a severity biomarker for severe COVID-19 useful to distinguish the worst diagnostic and a potential target for innovative therapeutic strategies lead to a direct treatment for severe COVID-19.

## Supporting information

Uncut gels

## Data Availability

All data produced in the present work are contained in the manuscript.

## ACKNOWLEDGEMENTS

This work was supported by grants from CNPq (311905/2019-6) and FAPERJ (E-26/210.240/2020; E-26/210.823/2021; E-26/202.834/2018). We greatly acknowledge Marcia Amorim to participate in this study collecting blood from the participants of healthy controls, Daniele Cristina Passos da Rocha to perform the initial step of HMGB1 ELISA and Dr. Marcelo Nunes Afonso Teixeira and Dr. Fernanda Cruz to help us to interpret the medical records of the patients.

## AUTHORS CONTRIBUTIONS

A.R.R.V., C.C. and C.F.B. wrote the manuscript. A.R.R.V., C.C., R.V.J. and C.F.B. designed the research. A.R.R.V., C.C., C.F.B., V.S.F.J., M.P., A.C.N.B., N.R.A., L.E.B.S., S.P.C.B., S.N.F. and D.R. performed the research. A.R.R.V., C.C., C.F.B., V.S.F.J., M.P., A.C.N.B., N.R.A., L.E.B.S., S.P.C.B., S.N.F. A.C.O., J.S. and D.A. analyzed the data. D.A., M.R.F., J.S., A.M.V., A.C.O., R.C.S. and L.E.B.S. contributed new reagents/analytical tools. C.F.B. granted and supervised the team.

---

**Supp Fig 1. Spearman’s correlation analyses between inflammatory mediators and tissue injury biomarkers with HMGB1. (A-I)** Spearman’s correlation analyses between HMGB1 serum levels versus ATP, TF, PGE_2_, LTB_4_, creatinine, PO_2_/FiO_2_, lactate, CRP, and LDH levels. We plotted the highest value obtained for HMGB1 after analysis of two or three samples received from each COVID-19 patient. The values for ATP, TF, PGE2, LTB4, creatinine, PO_2_/FiO_2_, lactate, CRP, and LDH were paired exactly with the same sample chosen for analysis of HMGB1. No correlations were detected with the analyzed parameters.

**Uncut gels. Representative western blot for tissue factor and ferritin for serum samples of the subjects**. The samples were run blindly. The graph in the figure 2-6, panel C was obtained after the densitometry analysis of the western blot. In blue patients who discharged, in red patients who deceased, in green healthy controls and in black patients without medical records and excluded to the cohort.

## REFERENCES

1. Schultze JL, Aschenbrenner AC. COVID-19 and the human innate immune system. Cell. 2021;184(7):1671–1692. doi:10.1016/j.cell.2021.02.029

2. Sokolowska M, Lukasik ZM, Agache I, et al. Immunology of COVID[19: Mechanisms, clinical outcome, diagnostics, and perspectives—A report of the European Academy of Allergy and Clinical Immunology (EAACI). Allergy. 2020;75(10):2445–2476. doi:10.1111/all.14462

3. Willi S, Lüthold R, Hunt A, et al. COVID-19 sequelae in adults aged less than 50 years: A systematic review. Travel Med Infect Dis. 2021;40:101995. doi:10.1016/j.tmaid.2021.101995

4. Bellan M, Soddu D, Balbo PE, et al. Respiratory and Psychophysical Sequelae Among Patients With COVID-19 Four Months After Hospital Discharge. JAMA Netw Open. 2021;4(1):e2036142. doi:10.1001/jamanetworkopen.2020.36142

5. Mitrani RD, Dabas N, Goldberger JJ. COVID-19 cardiac injury: Implications for long-term surveillance and outcomes in survivors. Hear Rhythm. 2020;17(11):1984–1990. doi:10.1016/j.hrthm.2020.06.026

6. sahin ahmet riza. Multisystemic Long-Term Sequelae of Covid-19: A Review Based on the Current Literature Over a Year of Pandemic Experience. Eurasian J Med Oncol. Published online 2021. doi:10.14744/ejmo.2021.69960

7. Wang F, Kream RM, Stefano GB. Long-Term Respiratory and Neurological Sequelae of COVID-19. Med Sci Monit. 2020;26. doi:10.12659/MSM.928996

8. Wang J, Jiang M, Chen X, Montaner LJ. Cytokine storm and leukocyte changes in mild versus severe SARS[CoV[2 infection: Review of 3939 COVID[19 patients in China and emerging pathogenesis and therapy concepts. J Leukoc Biol. 2020;108(1):17–41. doi:10.1002/JLB.3COVR0520-272R

9. Bonaldi T. Monocytic cells hyperacetylate chromatin protein HMGB1 to redirect it towards secretion. EMBO J. 2003;22(20):5551–5560. doi:10.1093/emboj/cdg516

10. Scaffidi P, Misteli T, Bianchi ME. Release of chromatin protein HMGB1 by necrotic cells triggers inflammation. Nature. 2002;418(6894):191–195. doi:10.1038/nature00858

11. Chi W, Chen H, Li F, Zhu Y, Yin W, Zhuo Y. HMGB1 promotes the activation of NLRP3 and caspase-8 inflammasomes via NF-κB pathway in acute glaucoma. J Neuroinflammation. 2015;12(1):137. doi:10.1186/s12974-015-0360-2

12. Deng M, Tang Y, Li W, et al. The Endotoxin Delivery Protein HMGB1 Mediates Caspase-11-Dependent Lethality in Sepsis. Immunity. 2018;49(4):740–753.e7. doi:10.1016/j.immuni.2018.08.016

13. Liu T, Barrett NA, Kanaoka Y, et al. Cysteinyl Leukotriene Receptor 2 (CysLT2R) Drives Lung Immunopathology Through a High Mobility Box 1-Dependent Mechanism. J Allergy Clin Immunol. 2018;141(2):AB206. doi:10.1016/j.jaci.2017.12.650

14. Liu T, Barrett NA, Kanaoka Y, et al. Cysteinyl leukotriene receptor 2 drives lung immunopathology through a platelet and high mobility box 1-dependent mechanism. Mucosal Immunol. 2019;12(3):679–690. doi:10.1038/s41385-019-0134-8

15. Andersson U, Yang H, Harris H. High-mobility group box 1 protein (HMGB1) operates as an alarmin outside as well as inside cells. Semin Immunol. 2018;38:40–48. doi:10.1016/j.smim.2018.02.011

16. Chen L, Long X, Xu Q, et al. Elevated serum levels of S100A8/A9 and HMGB1 at hospital admission are correlated with inferior clinical outcomes in COVID-19 patients. Cell Mol Immunol. 2020;17(9):992–994. doi:10.1038/s41423-020-0492-x

17. Chen R, Huang Y, Quan J, et al. HMGB1 as a potential biomarker and therapeutic target for severe COVID-19. Heliyon. 2020;6(12):e05672. doi:10.1016/j.heliyon.2020.e05672

18. Sivakorn C, Dechsanga J, Jamjumrus L, et al. High Mobility Group Box 1 and Interleukin 6 at Intensive Care Unit Admission as Biomarkers in Critically Ill COVID-19 Patients. Am J Trop Med Hyg. 2021;105(1):73–80. doi:10.4269/ajtmh.21-0165

19. Wei J, Alfajaro MM, DeWeirdt PC, et al. Genome-wide CRISPR Screens Reveal Host Factors Critical for SARS-CoV-2 Infection. Cell. 2021;184(1):76–91.e13. doi:10.1016/j.cell.2020.10.028

20. Siddiqi HK, Mehra MR. COVID-19 illness in native and immunosuppressed states: A clinical–therapeutic staging proposal. J Hear Lung Transplant. 2020;39(5):405–407. doi:10.1016/j.healun.2020.03.012

21. Lin Y, Wu Y, Zhong P, et al. A clinical staging proposal of the disease course over time in non-severe patients with coronavirus disease 2019. Sci Rep. 2021;11(1):10681. doi:10.1038/s41598-021-90111-y

22. Dalli J, Colas RA, Quintana C, et al. Human Sepsis Eicosanoid and Proresolving Lipid Mediator Temporal Profiles. Crit Care Med. 2017;45(1):58–68. doi:10.1097/CCM.0000000000002014

23. Zaid Y, Doré É, Dubuc I, et al. Chemokines and eicosanoids fuel the hyperinflammation within the lungs of patients with severe COVID-19. J Allergy Clin Immunol. 2021;148(2):368–380.e3. doi:10.1016/j.jaci.2021.05.032

24. Davis HE, Assaf GS, McCorkell L, et al. Characterizing long COVID in an international cohort: 7 months of symptoms and their impact. EClinicalMedicine. 2021;38:101019. doi:10.1016/j.eclinm.2021.101019

25. Ryan FJ, Hope CM, Masavuli MG, et al. Long-term perturbation of the peripheral immune system months after SARS-CoV-2 infection. BMC Med. 2022;20(1):26. doi:10.1186/s12916-021-02228-6

26. McKechnie JL, Blish CA. The Innate Immune System: Fighting on the Front Lines or Fanning the Flames of COVID-19? Cell Host Microbe. 2020;27(6):863–869. doi:10.1016/j.chom.2020.05.009

27. Dorneles GP, Teixeira PC, da Silva IM, et al. Alterations in CD39/CD73 Axis of T cells associated with COVID-19 severity. [Preprinted].:2021.09.18.21263782. doi:10.1101/2021.09.18.21263782. Posted on medRxiv October 15, 2021.

28. Marinho Y, Marques-da-Silva C, Santana PT, et al. MSU Crystals induce sterile IL-1β secretion via P2X7 receptor activation and HMGB1 release. Biochim Biophys Acta - Gen Subj. 2020;1864(1):129461. doi:10.1016/j.bbagen.2019.129461

29. Toki Y, Takenouchi T, Harada H, et al. Extracellular ATP induces P2X7 receptor activation in mouse Kupffer cells, leading to release of IL-1β, HMGB1, and PGE2, decreased MHC class I expression and necrotic cell death. Biochem Biophys Res Commun. 2015;458(4):771–776. doi:10.1016/j.bbrc.2015.02.011

30. Leclerc P, Wähämaa H, Idborg H, Jakobsson PJ, Harris HE, Korotkova M. IL[1β/HMGB1 Complexes Promote The PGE 2 Biosynthesis Pathway in Synovial Fibroblasts. Scand J Immunol. 2013;77(5):350–360. doi:10.1111/sji.12041

31. Jaulmes A, Thierry S, Janvier B, et al. Activation of sPLA2[IIA and PGE2 production by high mobility group protein B1 in vascular smooth muscle cells sensitized by IL[1β. FASEB J. 2006;20(10):1727–1729. doi:10.1096/fj.05-5514fje

32. Pérez MM, Pimentel VE, Fuzo CA, et al. Cholinergic and lipid mediators crosstalk in Covid-19 and the impact of glucocorticoid therapy. [Preprinted]. doi:10.1101/2021.01.07.20248970. Posted on medRxiv January 09, 2021.

33. García-Arnandis I, Guillén MI, Gomar F, Pelletier J-P, Martel-Pelletier J, Alcaraz MJ. High mobility group box 1 potentiates the pro-inflammatory effects of interleukin-1β in osteoarthritic synoviocytes. Arthritis Res Ther. 2010;12(4):R165. doi:10.1186/ar3124

34. Hreggvidsdottir HS, Östberg T, Wähämaa H, et al. The alarmin HMGB1 acts in synergy with endogenous and exogenous danger signals to promote inflammation. J Leukoc Biol. 2009;86(3):655–662. doi:10.1189/jlb.0908548

35. Harris HE, Andersson U, Pisetsky DS. HMGB1: A multifunctional alarmin driving autoimmune and inflammatory disease. Nat Rev Rheumatol. 2012;8(4):195–202. doi:10.1038/nrrheum.2011.222

36. Ahmadi P, Hartjen P, Kohsar M, et al. Defining the CD39/CD73 Axis in SARS-CoV-2 Infection: The CD73-Phenotype Identifies Polyfunctional Cytotoxic Lymphocytes. Cells. 2020;9(8):1750. doi:10.3390/cells9081750

37. Hottz ED, Azevedo-Quintanilha IG, Palhinha L, et al. Platelet activation and platelet-monocyte aggregate formation trigger tissue factor expression in patients with severe COVID-19. Blood. 2020;136(11):1330–1341. doi:10.1182/blood.2020007252

38. Vicentino ARR, Carneiro VC, Allonso D, et al. Emerging Role of HMGB1 in the Pathogenesis of Schistosomiasis Liver Fibrosis. Front Immunol. 2018;9:1979. doi:10.3389/fimmu.2018.01979

39. Varikasuvu SR, Varshney S, Dutt N, et al. D-dimer, disease severity, and deaths (3D-study) in patients with COVID-19: a systematic review and meta-analysis of 100 studies. Sci Rep. 2021;11(1):21888. doi:10.1038/s41598-021-01462-5

40. Ogawa EN, Ishizaka A, Tasaka S, et al. Contribution of High-Mobility Group Box-1 to the Development of Ventilator-induced Lung Injury. Am J Respir Crit Care Med. 2006;174(4):400–407. doi:10.1164/rccm.200605-699OC

41. Ito I, Fukazawa J, Yoshida M. Post-translational Methylation of High Mobility Group Box 1 (HMGB1) Causes Its Cytoplasmic Localization in Neutrophils. J Biol Chem. 2007;282(22):16336–16344. doi:10.1074/jbc.M608467200

42. Vogel S, Bodenstein R, Chen Q, et al. Platelet-derived HMGB1 is a critical mediator of thrombosis. J Clin Invest. 2015;125(12):4638–4654. doi:10.1172/JCI81660

43. Bianchi ME. HMGB1 loves company. J Leukoc Biol. 2009;86(3). doi:10.1189/jlb.1008585

44. Peng T, Du S-Y, Son M, Diamond B. HIF-1α is a negative regulator of interferon regulatory factors: Implications for interferon production by hypoxic monocytes. Proc Natl Acad Sci. 2021;118(26):e2106017118. doi:10.1073/pnas.2106017118

45. Moreira FRR, D’arc M, Mariani D, et al. Epidemiological dynamics of SARS-CoV-2 VOC Gamma in Rio de Janeiro, Brazil. Virus Evol. 2021;7(2). doi:10.1093/ve/veab087

46. Allonso D, Belgrano FS, Calzada N, Guzmán MG, Vázquez S, Mohana-Borges R. Elevated serum levels of high mobility group box 1 (HMGB1) protein in dengue-infected patients are associated with disease symptoms and secondary infection. J Clin Virol. 2012;55(3):214–219. doi:10.1016/j.jcv.2012.07.010

47. Alvim RGF, Lima TM, Rodrigues DAS, et al. Development and large-scale validation of a highly accurate SARS-COV-2 serological test using regular test strips for autonomous and affordable finger-prick sample collection, transportation, and storage. [Preprinted]. doi:10.1101/2020.07.13.20152884. Posted on medRxiv July 07, 2021

