## Supplementary figures and images for "HMGB1 correlates with severity and death of COVID-19 patients"

### Sup Fig 1

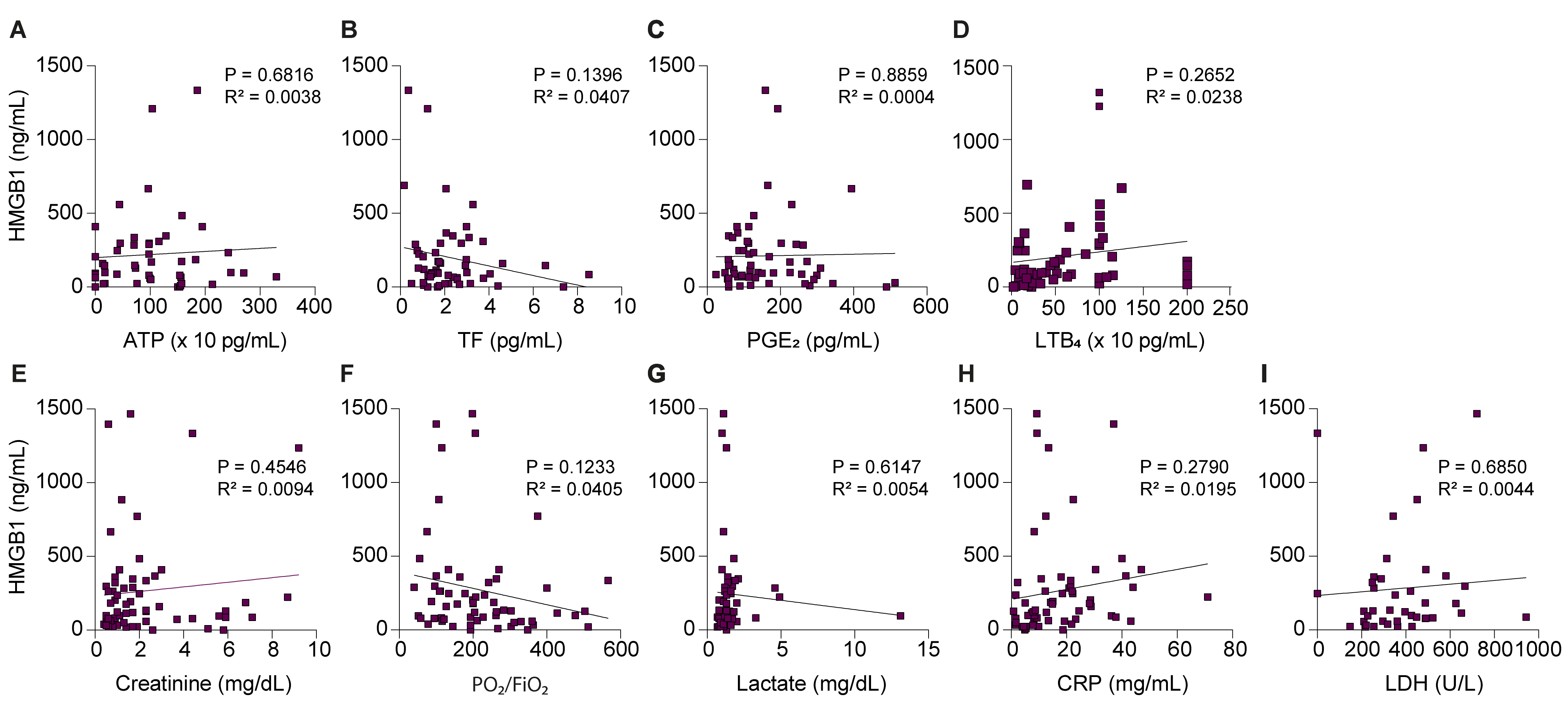
