## Supplementary material for "HMGB1 correlates with severity and death of COVID-19 patients": Uncut gels

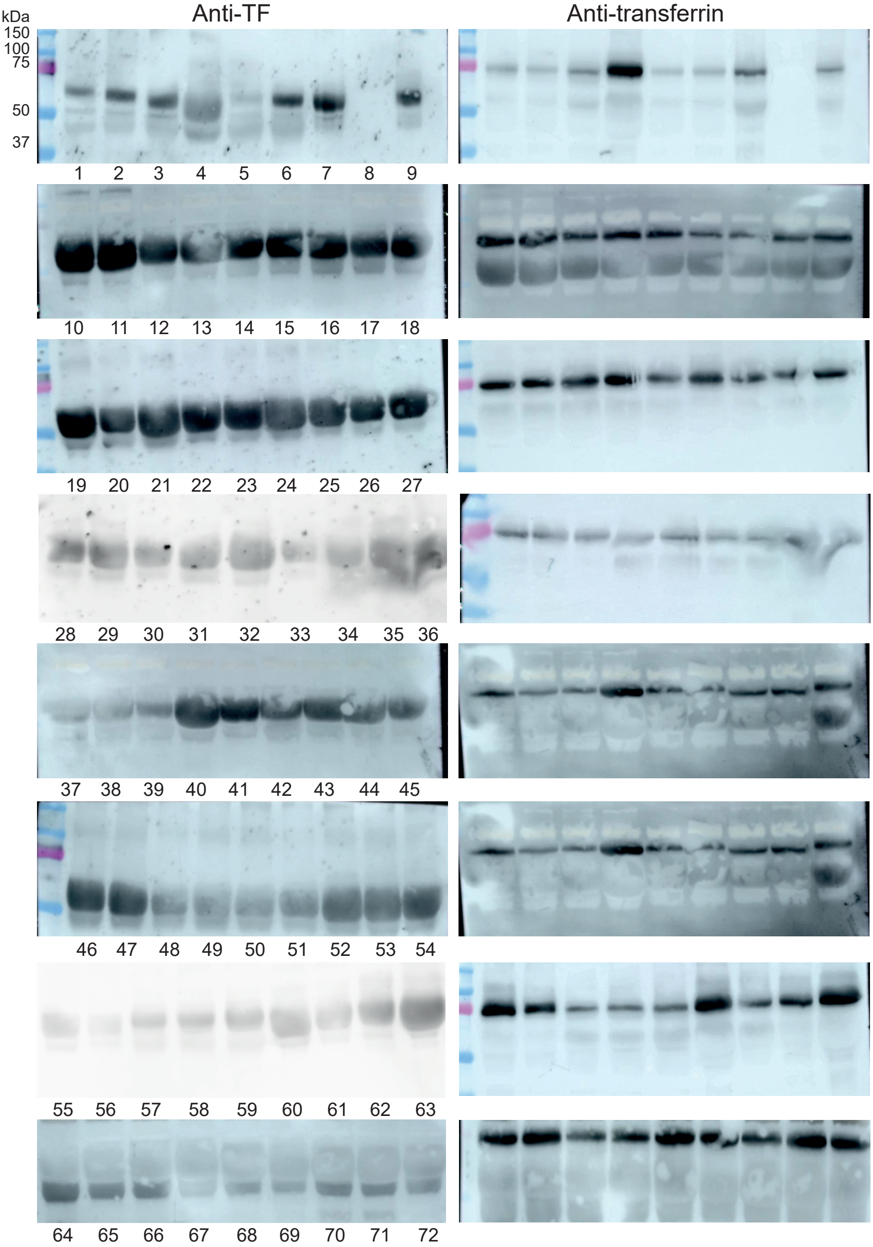


Samples legends: Healthy controls; Discharged; Deceased; Patients without medical records

1 - COV497, 1ST BLOOD COLLECTION (BC)
2 - COV497, 2ND BC
3 - COV497, 3RD BC
4 - COV528, 1ST BC
5 - COV528, 2ND BC
6 - COV528, 3RD BC
7 - COV613, 1ST BC
8 - COV613, 2ND BC
9 - COV613, 3RD BC
10 - COV1824, 1ST BC
11 - COV1824, 2ND BC
12 - COV1834, 1ST BC
13 - COV1834, 2ND BC
14 - COV1834, 3RD BC
15 - COV1859, 1ST BC
16 - COV1859, 2ND BC
17 - COV1859, 3RD BC
18 - COV1681
19 - COV4903, 1ST BC
20 - COV4903, 2ND BC
21 - COV5241, 1ST BC
22 - COV5241, 2ND BC
23 - COV5318, 1ST BC
24 - COV5318, 2ND BC
25 - COV5349, 1ST BC
26 - COV5349, 2ND BC
27 - COV5361, 1ST BC
28 - COV843, 1ST BC
29 - COV843, 2ND BC
30 - COV843, 3RD BC
31 - COV900, 1ST BC
32 - COV900, 2ND BC
33 - COV900, 3RD BC
34 - CTRL1
35 - CTRL4
36 - CTRL8
37 - COV347, 1ST BC
38 - COV347, 2ND BC
39 - COV409, 1ST BC
40 - COV471, 1ST BC
41 - COV471, 2ND BC
42 - COV518, 1ST BC
43 - COV1759, 1ST BC
44 - COV1759, 2ND BC
45 - COV1983
46 - COV2950, 1ST BC
47 - COV3010, 1ST BC
48 - COV3010, 2ND BC
49 - COV3066, 1ST BC
50 - COV3066, 2ND BC
51 - COV3112, 1ST BC
52 - COV3112, 1ST BC
53 - COV3433, 1ST BC
54 - COV3433, 2ND BC
55 - COV1471, 1ST BC
56 - COV1471, 2ND BC
57 - COV1471, 3RD BC
58 - COV1616, 1ST BC
59 - COV1616, 2ND BC
60 - COV1616, 3RD BC
61 - CTRL15
62 - CTRL23
63 - CTRL25
64 - COV347, 1ST BC
65 - COV347, 2ND BC
66 - COV409, 1ST BC
67 - COV471, 1ST BC
68 - COV471, 2ND BC
69 - COV518, 1ST BC
70 - COV1759, 1ST BC
71 - COV1759, 2ND BC
72 - COV1983

**Uncut gels: Representative western blot for tissue factor (TF) and transferrin for serum samples of the subjects.** The samples were run blindly. The graphs in the figure 2-6, panel C were obtained after the densitometry of each bands. In green, healthy controls. In blue, patients who discharged. In red, patients who deceased. In black, patients without medical records and were excluded from the cohort.
